# Automated large-scale AMD progression prediction using machine-read OCT biomarkers

**DOI:** 10.1101/2022.08.21.22278906

**Authors:** Akos Rudas, Jeffrey N. Chiang, Giulia Corradetti, Nadav Rakocz, Eran Halperin, Srinivas R. Sadda

**Affiliations:** Department of Computational Medicine, University of California - Los Angeles, Los Angeles, CA; Doheny Eye Institute, Pasadena, CA; Department of Ophthalmology, David Geffen School of Medicine, University of California-Los Angeles, Los Angeles, California, United States; Department of Computer Science, University of California - Los Angeles, Los Angeles, CA

**Author notes:** **Corresponding Author:** SriniVas R Sadda, Doheny Eye Institute, Department of Ophthalmology, David Geffen School of Medicine at UCLA, 150 N Orange Grove Blvd, Pasadena, CA 91103, USA. Last Authors equal contribution (Eran Halperin, Srinivas R. Sadda). **Prior submission to other journals:** None. **Ethical statement:** the authors are accountable for all aspects of the work in ensuring that questions related to the accuracy or integrity of any part of the work are appropriately investigated and resolved.

**Keywords:** Age-related Macular Degeneration, deep learning, exudative AMD, AMD biomarkers, Machine Learning

## Abstract

Age-related Macular Degeneration (AMD) is a major cause of irreversible vision loss in individuals over 55 years old in the United States. While anti-vascular growth factor injections can be used to treat macular neovascularization (MNV), there are currently no treatments available to halt or reverse geographic atrophy, which is the late-stage of nonneovascular AMD. There is a great interest in detecting early biomarkers associated with a higher risk for AMD progression in order to design early intervention clinical trials. The annotation of structural biomarkers on optical coherence tomography (OCT) B-scans is a laborious, complex and time-consuming process, and discrepancies between human graders can introduce variability into this assessment.

To address this issue, a deep-learning model (SLIVER-net) was proposed, which could identify AMD biomarkers on structural OCT volumes with high precision and without human supervision. However, the validation was performed on a small dataset, and the true predictive power of these detected biomarkers in the context of a large cohort has not been evaluated. In this retrospective cohort study, we perform the largest-scale validation of these biomarkers to date. We also assess how these features combined with other EHR data (demographics, comorbidities, etc) affect and/or improve the prediction performance relative to known factors. Our hypothesis is that these biomarkers can be identified by a machine learning algorithm without human supervision, in a way that they preserve their predictive nature.

The way we test this hypothesis is by building several machine learning models utilizing these machine-read biomarkers, and assessing their added predictive power. We found that not only can we show that the machine-read OCT B-scan biomarkers are predictive of AMD progression, we also observe that our proposed combined OCT and EHR data-based algorithm significantly outperforms the state-of-the-art solution in clinically relevant metrics and provides actionable information which has the potential to improve patient care. In addition, it provides a framework for automated large-scale processing of OCT volumes, making it possible to analyze vast archives without human supervision.

## Introduction

Age-related Macular Degeneration (AMD) represents the leading cause of irreversible blindness in subjects older than 55 years of age in developed countries.(Wong et al. 2014) As the population ages and life expectancy increases, the incidence of the disease is projected to rise.(Jonas et al. 2017) The late stage of the disease is characterized by the presence of geographic atrophy (GA), macular atrophy (MA) or macular neovascularization (MNV).(Ferris et al. 2013) (Spaide et al. 2020; Sadda et al. 2018) Atrophic changes, such as GA or MA are associated with loss of the retinal pigment epithelium (RPE), choriocapillaris impairment, and photoreceptor degeneration leading to irreversible vision loss. (Blair 1975) No proven therapies are currently available for the late stage atrophic form of AMD, although many agents are under investigation.(Halawa et al. 2021) Micronutrient supplementation using the AREDS2 (Age-related Eye Disease Study) formulation has been shown to reduce progression from intermediate AMD to neovascular AMD, but there was no benefit in reducing the development of atrophy.(Age-Related Eye Disease Study 2 (ARED…; AREDS2 Research Group et al. 2012)

In contrast to atrophic AMD, anti-vascular endothelial growth factor (anti-VEGF) therapy has proven to be effective at reducing vision loss and even improving vision in eyes with neovascular or wet AMD. However, even with consistent treatment, vision loss and progression to atrophy may occur even in eyes with MNV.(Chandra et al. 2020) (Daniel et al. 2014) Studies have shown that best visual outcomes are achieved by detecting the neovascular disease activity early and treating before significant visual loss has occurred. (Kodjikian et al. 2021) (Yoshida et al. 2020) (Dervenis and Younis 2016)

As a result of this desire to detect disease progression early on, there has been significant effort to identify biomarkers which may predict the development of advanced AMD. Identification of biomarkers has been facilitated by the broad availability of optical coherence tomography (OCT), which has become the dominant imaging technology in ophthalmic clinical practice. Studies evaluating OCT have identified a number of features including high central drusen volume (hcDV), subretinal drusenoid deposits (SDD) and, or reticular pseudodrusen (RPD), intraretinal hyperreflective foci (IHRF), and hyporeflective drusen cores (hDC), which have been shown to be associated with a higher risk for progression to advanced AMD (Lei et al. 2017) (Corradetti et al. 2021; Nassisi et al. 2019). However, identification of these biomarkers requires extensive training and careful examination of the individual B-scans in the OCT volume – this may be challenging in the context of a busy clinical practice and may be susceptible to variability in interpretation among clinicians. Therefore, machine learning algorithms have been developed to automatically detect structural OCT B-scan biomarkers predictive for progression to advanced AMD. (SLIVER-net, Rakocz et al. 2021) By automating the interpretation of OCT volumes, this approach enables low-cost, large-scale studies and analyses of AMD progression while anchoring inferences and conclusions to clinically-relevant biomarkers. However, machine learning approaches in detecting early biomarkers of disease have been only tested in small cohorts, not accounting for heterogeneity in the prediction of the outcome between different environments, settings and populations. (Thakoor et al. 2022; de Jong et al. 2021)

In the present study, we offer the largest machine learning validation to date of these structural OCT B-scan biomarkers predictive for AMD progression. Our hypothesis is that these biomarkers can be inferred by a machine learning algorithm without human supervision, in a way that they preserve their predictive nature. The way we test this hypothesis is by building machine learning models upon these machine-read biomarkers, and assess their predictive power. Consequently, we also validate the high accuracy with which SLIVER-net automatically detects structural OCT B-scan biomarkers in a large cohort. Our model is not only capable of successfully detecting these structural OCT biomarkers, but also able to predict future AMD progression and prognosis, which may impact clinical decision making. First, we explore the ability of the automated approach to *predict* future conversion to exudative AMD within 2 years from the baseline OCT. Then, we apply our approach to diagnosis, showing that machine-read OCT features are also informative for determining the current disease status. Our approach is able to significantly improve predictive models which consider only the currently available risk factors, and are developed using data from smaller cohorts with less population heterogeneity.

## Results

Machine-read OCT features were evaluated for their clinical utility relative to currently known risk factors contained within the electronic health record using a predictive modeling framework. These features were evaluated in their ability to predict conversion to exudative AMD as well as diagnosis of current exudative AMD.

### Predicting future conversion to Exudative AMD

Using machine-read OCT B-scans features and EHR-derived risk factors together in logistic regression models (*combined*), we were able to successfully predict exudative AMD conversion within two years with an area under the ROC curve (AUROC) of 0.82 (95% confidence interval (CI): 0.78, 0.85) and area under the Precision Recall Curve (AUPRC) of 0.49 (95% CI:0.41, 0.57).

Relative to the EHR-derived features of age, sex, race, smoking status, and comorbidities, the addition of machine-read OCT B-scans features resulted in significantly improved predictive performance in terms of AUROC and AUPRC (see Figure 1). The trivial model (*current status*) utilizing only the presence of dry AMD at the time of examination and the time to the next examination, yielded an AUROC of 0.57 (95% CI: 0.54, 0.60) and AUPRC of 0.21 (95% CI: 0.18, 0.24). With added EHR-derived features and comorbidities (*risk factors*), the performance increased to AUROC of 0.72 (95% CI: 0.69, 0.74) and AUPRC of 0.25 (95% CI: 0.22, 0.28). The machine-read OCT B-scan features (*biomarkers*) were also by themselves highly predictive of exudative AMD conversion (Figures 1, 2; biomarkers) yielding AUROC of 0.80 (0.78, 0.82) and AUPRC of 0.46 (0.41, 0.50).

**Figure 1.**
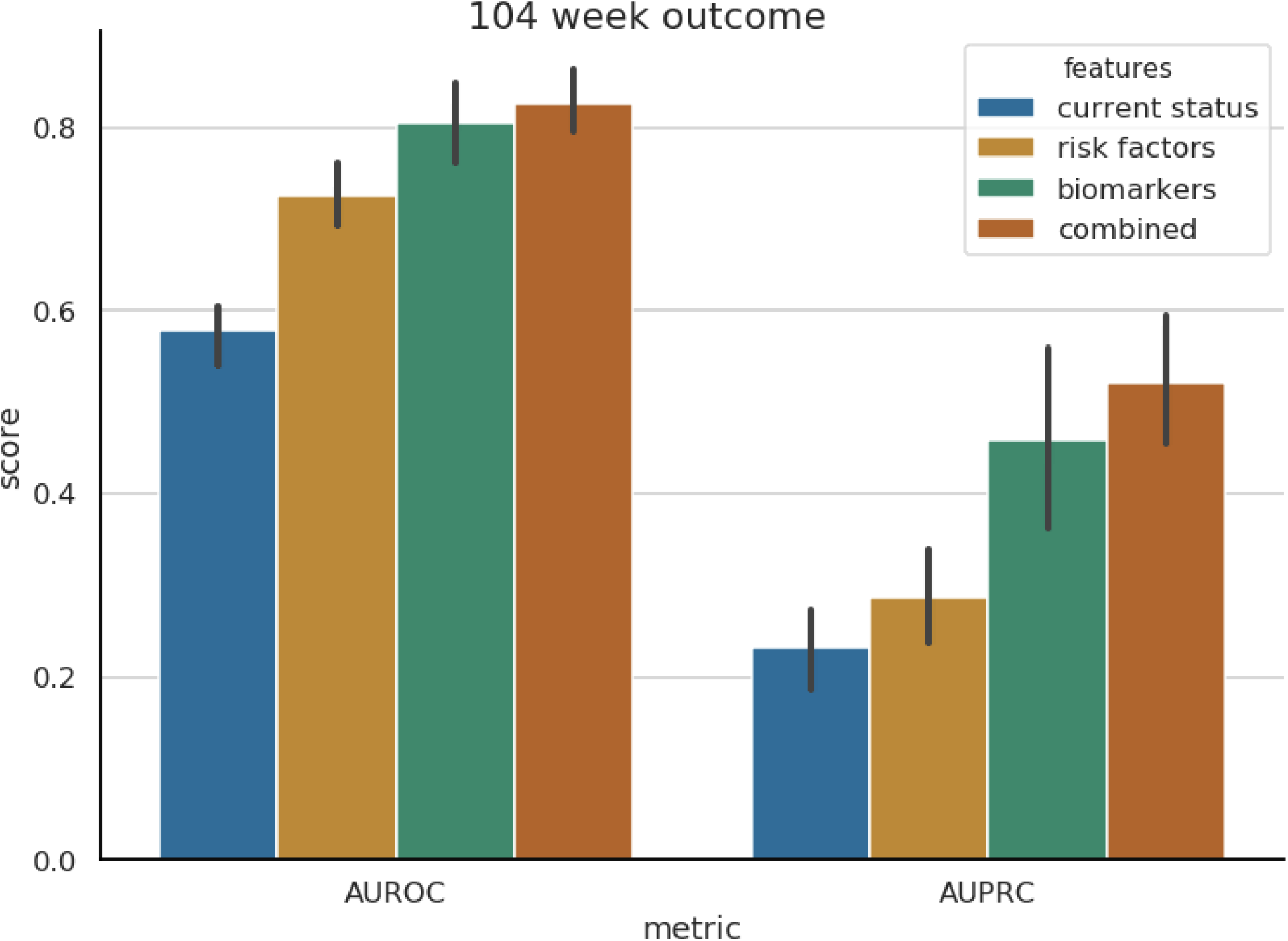
Exudative AMD prediction performance. Left: Areas Under the Receiver Operating Characteristic (ROC) curve. Right: Areas Under the Precision-Recall (PR) curve. Each bar represents the performance utilizing a different set of features (see legend). Error lines represent the 95% confidence intervals, computed using bootstrapping.

**Figure 2.**
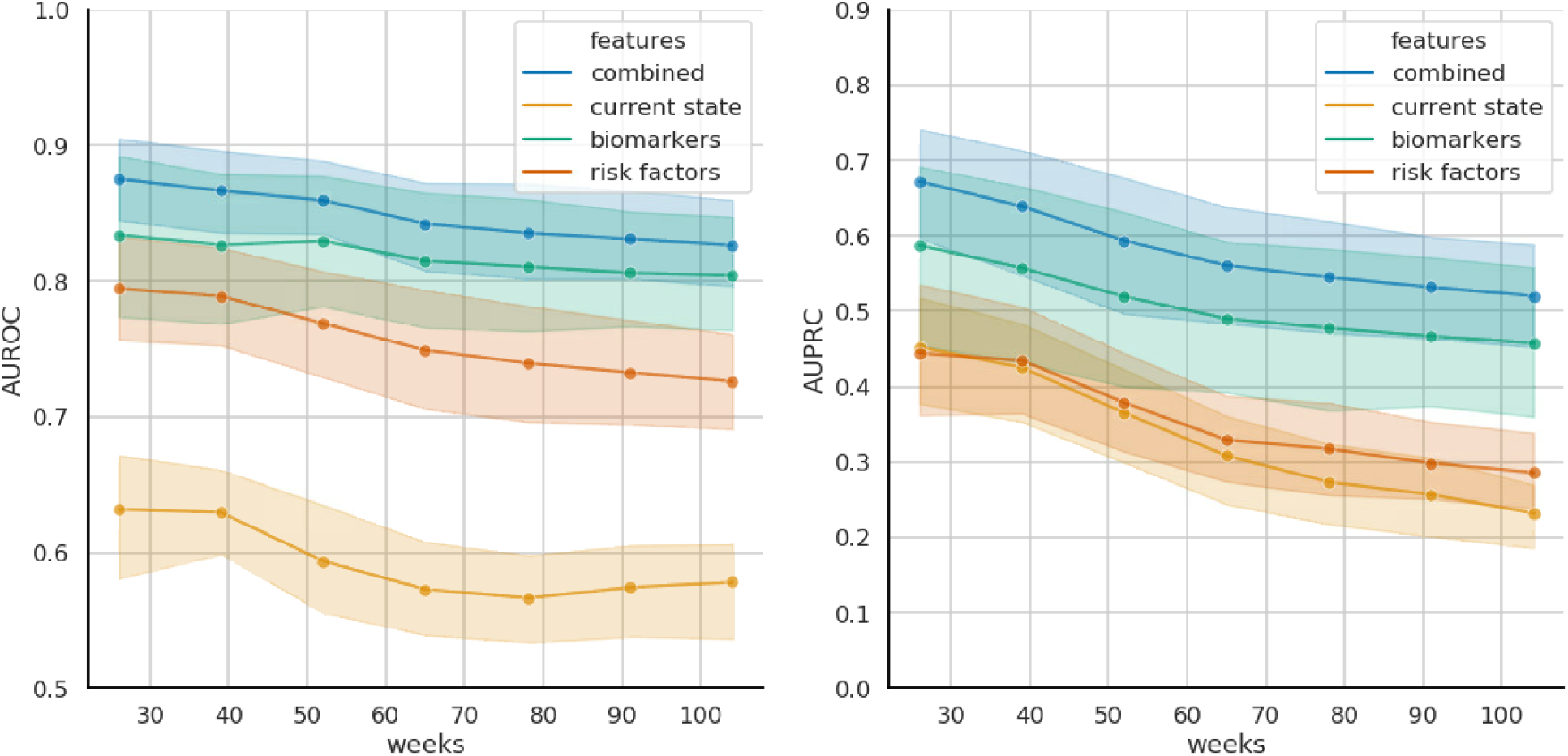
Prediction of exudative AMD Conversion. L*eft*. Area under the ROC curve (AUROC) as a function of prediction time frame. *Right*. Area under the Precision-Recall curve (AUPRC) as a function of prediction time frame. 95% Confidence intervals were computed using bootstrapping.

Patients could have converted to exudative AMD at any point during the two year window evaluated. To observe predictive performance over time, the above analysis was repeated at 3 month intervals. When additional models were trained for their ability to predict exudative AMD conversion within 3, 6, 9, 12, 15, 18, 21, and 24 months, up to within two years, we observed the general trend that AUROC was statistically stable across time periods, while there was a general decrease in AUPRC (Figure 2). The presence of the biomarkers appeared to be more indicative of imminent exudative AMD conversion.

Recent work has also applied deep learning to raw OCT volumes to predict 6-month wet AMD conversion in the fellow eye when a patient already had wet AMD in one eye (Yim et al. 2020). In the fellow eye of patients who already had exudative AMD, we performed a post-hoc analysis on model performance.

Table 2 reports a detailed view of all performance metrics for the full model at different operating thresholds for 26 and 104 weeks. Table 3 shows a more detailed comparison with different operating thresholds and other performance metrics from (Yim et al. 2020) using their selection criteria.

**Table 2.**
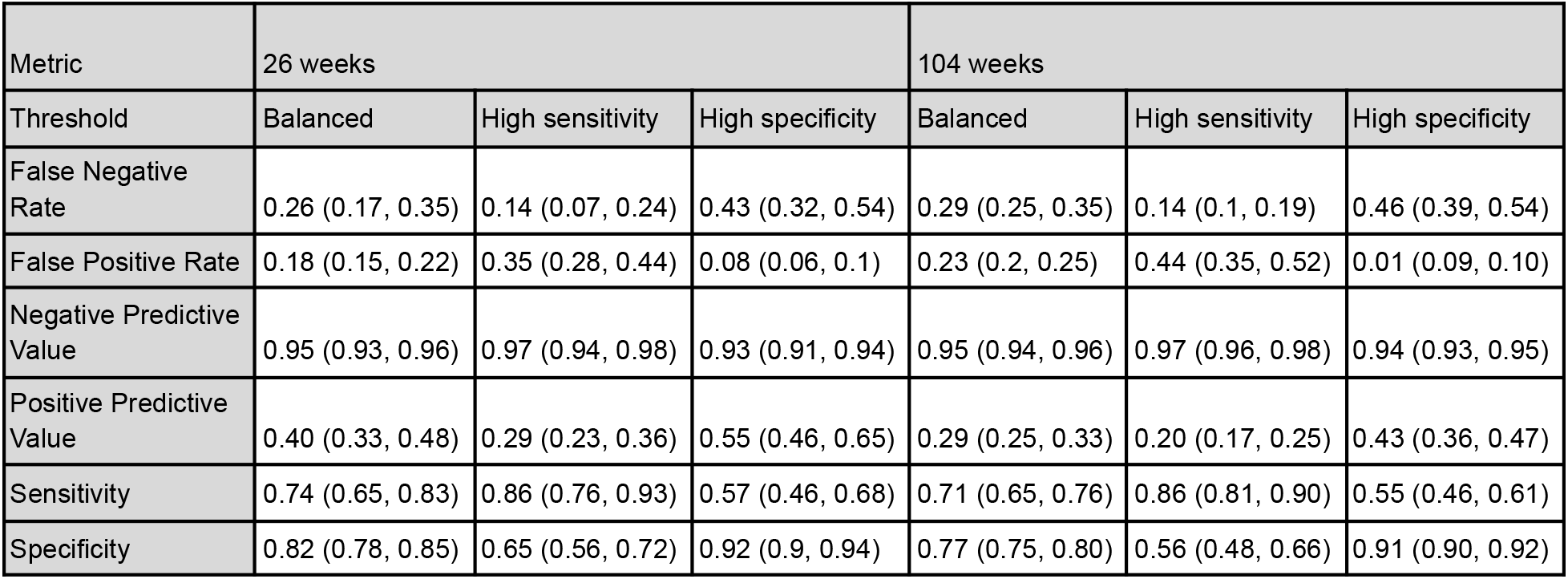
Performance metrics of the combined prediction model for a timeframe of 26 and 104 weeks. Results using a threshold selected for high sensitivity (>80%), a threshold for high specificity (>90%), and one for a balanced case are presented.

**Table 3.**
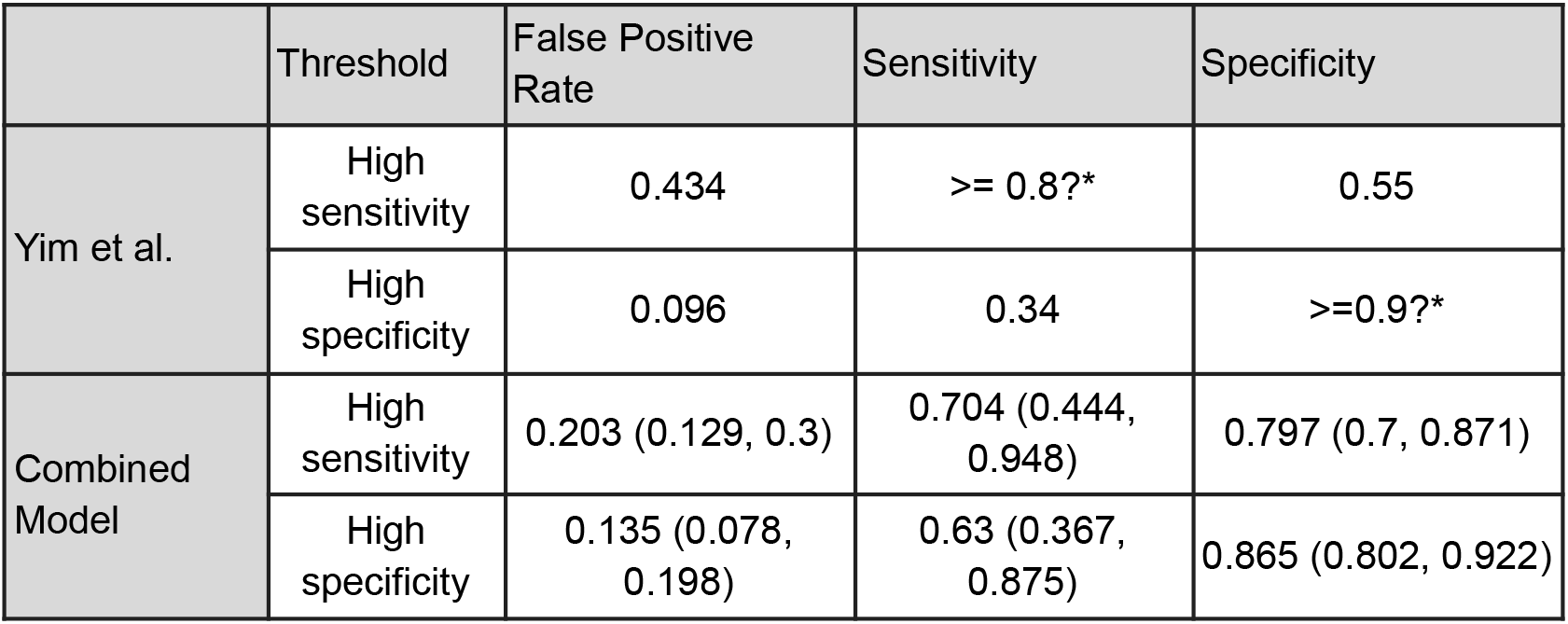
Performance metric comparison of our combined model and reported performance metrics of the model proposed by (Yim et al. 2020) using the same selection criteria. *Question marks indicate values not clearly reported.

### Large scale validation of machine-read OCT features for diagnosis

Although these structural OCT B-scan biomarkers are expected to be predictors of AMD progression, and not biomarkers to base a diagnosis on, as an experiment, we applied the same logistic regression framework in order to diagnose the current eye with exudative AMD. EHR and machine-read OCT B-scan features were used as input features to diagnose exudative AMD. We observed that relative to the EHR-derived features of age, sex, race, smoking status, and comorbidities, which achieved diagnostic performance of AUROC 0.82 (95% CI: 0.81, 0.83) and AUPRC 0.34 (95% CI: 0.32, 0.37), the addition of machine-read OCT B-scan features resulted in significantly improved diagnostic performance in terms of both AUROC [0.91 (95% CI: 0.90, 0.92)] and AUPRC [0.53 (95% CI: 0.50, 0.56)] (Figure 3). This improvement, based on the addition of machine-read OCT B-scan features, was consistent with a clinically validated scoring system (Lei et al. 2017), in which the presence of SDD, IHRF, and hcDV were associated with higher disease severity and progression.

**Figure 3.**
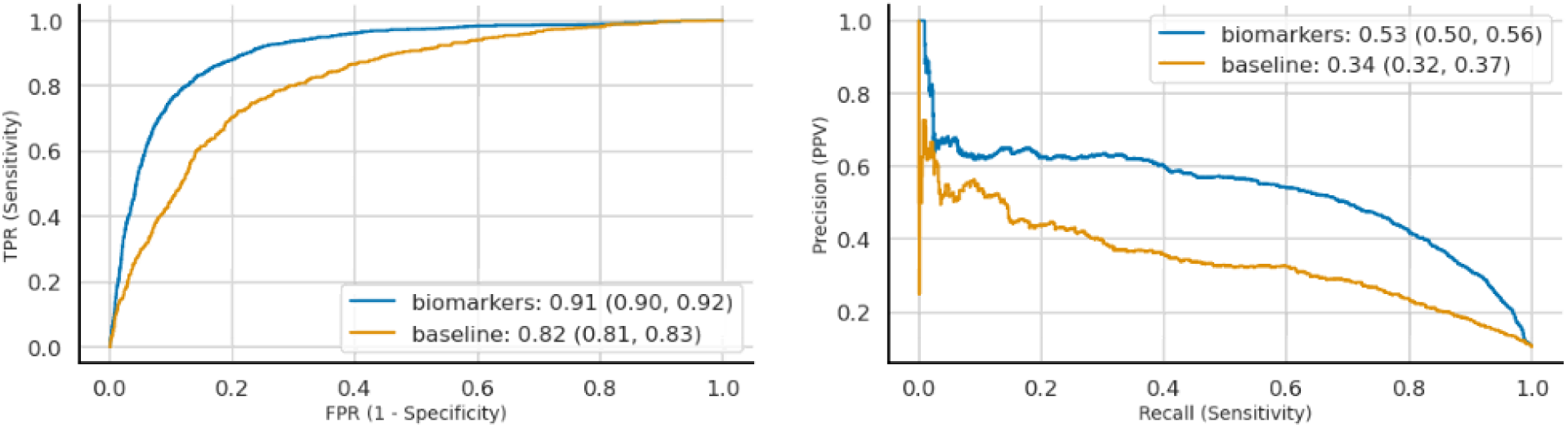
Automated diagnosis of exudative AMD. Curves correspond to models trained on different feature sets (see legend). *Left*. Receiver Operating Characteristics (ROC). *Right*. Precision-Recall curve (PRC). 95% Confidence intervals were computed using bootstrapping. Baseline model utilizes EHR-derived features and risk factors, the biomarker model includes machine-read OCT biomarkers too.

## Discussion

In this study we provide the first large-scale validation of machine-read structural OCT B-scan biomarkers for AMD progression. We do so by utilizing a deep learning method, SLIVER-net, which was trained to identify these biomarkers from OCT volumes. We show that regression models using these biomarkers do indeed predict AMD progression, thus validating not only the accuracy of SLIVER-net, but also the generalizability of these previously proposed structural OCT B-scan biomarkers, and the fact that they can be accurately inferred by a machine learning algorithm without human supervision. The prediction was implemented using a cross validation approach across 15000 OCT volumes collected from nearly 4200 patients.

To validate the utility of machine-read OCT B-scan biomarkers, we automatically predicted conversion to exudative AMD from nonexudative AMD. The automated assessment of conversion to exudative AMD was based on EHR data and OCT B scans volume data from subjects evaluated at ophthalmology clinics affiliated with a large academic hospital during 2018. The outcome (conversion to exudative AMD) was explored using several models and considering the following covariates: current AMD status, EHR-derived risk factors and comorbidities, and structural OCT B-scan biomarkers for progression of AMD. The ability of logistic regression models, trained to predict future conversion to exudative AMD, improved when adding AMD progression biomarkers to comorbidity features and demographic risk factors. Within a minimum of 3 months to a maximum of 2 years, logistic regression models trained with machine-read biomarkers performed with an AUROC of 0.82 (95% CI: 0.78, 0.85) and AUPRC of 0.49 (95% CI:0.41,0.57). This validation approach not only proved to be successful, but provided us with a clinically useful approach, which offers an early warning for the subset of patients identified as having a higher risk of AMD progression. Particularly, our study was performed on 4182 patients, while the largest study to date validating these biomarkers (Nassisi et al. 2019) included only 501 patients. De Fauw et al published about the ability of a deep learning algorithm to identify referral-warranted retinal diseases using structural OCT volumes from real-world practices, with a performance similar to human experts. (De Fauw et al. 2018) Although in this study the authors used (De Fauw et al. 2018) a dataset larger (N = 7,621) than ours, an important distinction is that our dataset included clinically more relevant annotations, using the AMD-related high risk biomarkers. We observed that diagnosis based on these biomarkers outperforms previously reported performance by deep learning approaches.

The detection of these biomarkers on structural OCT volumes requires human experts to manually perform the annotations or determinations, which can lead to measurement biases, inter-grader variability and a process which can be laborious and time-consuming. For these reasons, in real-world ophthalmic practice, the assessment of these biomarkers on OCT volumes is not yet part of the usual routine in clinical practice. Therefore, the validation of automated OCT annotations using machine learning algorithms is beneficial with the purpose to validate structural OCT features associated with a high-risk for progression to advanced AMD.

Exudation in eyes with macular neovascularization secondary to AMD appears in eyes with the late stage of the disease. The detection of fluid (exudation) at different levels within the retina (intraretinal, subretinal, sub-retinal pigment epithelium) defines the presence of disease activity. The advances in retinal imaging and the introduction of OCT technology have been transformative in the diagnosis, management and follow-up of eyes with exudative AMD, allowing the detection of fluid with high resolution and high precision. Of note, the exudative form of AMD can be successfully treated with anti-VEGF therapy, (Spaide et al. 2020) and it has been established that earlier treatment is associated with better visual outcomes (Schmidt-Erfurth et al. 2014). Therefore, there has been increasing interest in intervening at earlier stages of the disease. A number of studies have identified several high-risk biomarkers on structural OCT B-scan, such as intraretinal hyperreflective foci, subretinal drusenoid deposits, drusen with hyporeflective cores and high central drusen volume, which appear to be associated with a higher risk of progression from intermediate to late AMD.(Nassisi et al. 2019) (Lei et al. 2017; Nassisi et al. 2018) (Corradetti et al. 2021) (Nassisi et al. 2019) (Amarasekera et al. 2021) Our group has previously investigated the utility of SLIVER-net in automated detection of these high-risk biomarkers in a small annotated OCT dataset with good performance, sometimes better than retina specialists.(Rakocz et al. 2021) In this study, we used these AMD progression biomarkers to predict the conversion to exudative AMD.

For the scenario of predicting deterioration in 6 months, (Yim et al. 2020) reported an AUROC of 0.745 and an AUPRC of 0.123 on their test set. Our model reached a mean AUROC of 0.847 (0.716, 0.98) and mean AUPRC of 0.745 (0.539, 0.951) using the same cohort inclusion and exclusion criteria on our dataset. When binarizing their predictions to optimize for a high specificity around 90%, their model reportedly achieved 34% sensitivity and 9.6% false positive rate. Our model, which utilizes machine-read OCT B-scan biomarkers, yielded a 63% (36.7%, 87.5%) sensitivity and a13.5% (7.8%, 19.8%) false positive rate along the same optimization approach. They did not report positive predictive value (PPV) for the model, but in their paper, they included PPV metrics for three retinal specialists and three optometrists. Our model performed on par with them - every clinician’s performance (lowest reported: 18%; highest reported: 36.5% (Yim et al. 2020)) was within or below our model’s confidence intervals (Table 2, ‘26 weeks’). Our proposed model significantly outperformed the previous algorithm using similar data inclusion criteria (Supplementary Table 4).

We found that the machine-driven annotation was able to accurately predict the onset of exudative AMD within two years from the “baseline visit” at which the OCT was acquired. The importance of predicting the conversion to exudative AMD within 2 years is that it can impact the development of follow-up and monitoring schedules for a patient and for potentially selecting a higher-risk group of patients who may benefit from more expensive home-monitoring strategies.(Busquets and Sabbagh 2021) (Ho et al. 2021) (Pershing and Stein 2017) A personalized monitoring approach could potentially allow earlier detection of these patients, thereby leading to earlier therapeutic intervention and better visual outcomes.

Furthermore, while the SLIVER-net OCT biomarkers were clearly of additional predictive value, the EHR data alone provided significant predictive capability. This may be of relevance in resource-constrained countries where regular OCT imaging may not be feasible or available.

We note that our study has its limitations. Specifically, patients represented in our dataset visited ophthalmic clinics due to a scheduled check-up or an existing complaint or condition, and thus the selection of the patients may affect the generalizability of these results to the general population. However, since our work concentrates on the validation of established biomarkers that have been shown to be predictive of AMD progression in similar datasets (Lei et al. 2017; Nassisi et al. 2019), we do not expect this limitation to be particularly problematic, however we note that additional replication studies would be useful to further validate the biomarkers in the future.

Our study also has a few strengths. First, the machine learning algorithms have been trained and tested on a large cohort. We have performed a large-scale automatic validation of these previously established biomarkers, validating not only the biomarkers, but their automatic identification as well. Furthermore, we have provided evidence that automatic detection of structural OCT B-scan biomarkers using machine learning can be of value in predicting exudative AMD. The algorithm has the ability to provide automated annotation of these biomarkers on OCT volumes with high precision and feasibility, avoiding the laborious manual inspection or annotation of all the OCT B-scans.

In conclusion, we demonstrate on a large dataset that a machine learning algorithm can automatically annotate OCT volumes with high-risk structural OCT B-scan biomarkers of AMD progression with high accuracy. These annotations can be used to predict conversion to exudative AMD in eyes with nonexudative AMD with good performance, providing an impactful example of how machine learning has the ability to enhance patient care.

## Methods

### Study Design and Dataset

The study was conducted in compliance with the Declaration of Helsinki and approved by the UCLA Institutional Review Board (IRB, Ocular Imaging Study; Doheny – UCLA Eye Centers). The dataset consisted of 14,615 OCT volumes collected from 4,182 patients at affiliated Ophthalmology clinics during 2018 and corresponding electronic health record data for these visits including demographics, AMD status, and comorbidities (see Table 1).

**Table 1:** Descriptive statistics of the dataset

|  | Patients | Eyes | OCT volumes |
| --- | --- | --- | --- |
| Total | 4,182 | 8,075 | 14,615 |
| Prediction dataset | 1807 | 2615 | 4915 |
| Current Wet AMD (%) | 462 (11.0%) | 636 (7.9%) | 1,486 (10.2%) |
| No Wet AMD w/n 2 years | 3,478 | 7,134 | 12,523 |
| New Wet AMD w/n 6mo | 161 | 203 | 350 |
| New Wet AMD w/n 12 mo | 203 | 250 | 456 |
| New Wet AMD w/n 18 mo | 225 | 281 | 535 |
| New Wet AMD w/n 24 mo | 242 | 302 | 606 |
| Age (SD) | 66.45 (16.81) | 79.10 (8.43) | 81.57 (10.25) |
| Female | 53.8% | 58.7% | 57.9% |
| Never Smoker | 65.0% | 71.9% | 54.1% |
| Current Smoker | 2.8% | 1.8% | 4.6% |
| Latinx | 8.9% | 8.6% | 6.7% |
| Asian | 11.2% | 16.7% | 9.0% |
| Black | 4.7% | 1.8% | 1.6% |
| White | 66.4% | 67.9% | 72.9% |

#### EHR-derived features and outcomes

AMD status, demographics, and comorbidities were extracted from the electronic health records.

For each eye and visit, AMD-subtypes were defined using the following ICD-10 codes: nonexudative (dry) AMD (H35.31XX), exudative (wet) AMD (H35.32XX). The demographic factors extracted were age, sex, race, ethnicity, smoking status (Roizenblatt et al. 2018). Comorbidities were defined using the CMS (Chronic Condition Warehouse): cardiac arrhythmias, chronic pulmonary disease, congestive heart failure, diabetes (uncomplicated), hypertension, liver disease, metastatic cancer, obesity, renal failure, rheumatoid arthritis, valvular disease. All these clinical and demographic data were treated as dichotomous variables (presence/absence).

#### Automated quantification of AMD-related biomarkers

SLIVER-net (Rakocz et al. 2021) was used to automatically annotate OCT B-scan volumes for the following machine-read structural OCT AMD risk-progression biomarkers: high central drusen volume (hcDV), subretinal drusenoid deposits (SDD) and, or reticular pseudodrusen (RPD), intraretinal hyperreflective foci (IHRF), and hyporeflective drusen cores (hDC). The likelihood of each biomarker being present was represented as a score between 0 and 1. OCT B-scan volumes which we could not link to the EHR were not included in this analysis.

### Analyses

We used an 8-fold out-of-sample prediction framework in order to evaluate the predictive utility of the machine-read OCT biomarkers relative to EHR-derived features and risk factors for two tasks: 1) predicting conversion to future exudative AMD, and 2) diagnosis of current exudative AMD. We constructed several candidate feature sets, consisting of machine-read OCT and EHR-derived features and compared prediction performance for models trained using the different feature sets. All analyses were performed using Python, particularly the Scikit-learn(Pedregosa et al. 2011) and Statsmodels(Seabold and Perktold 2010) packages.

#### Predicting future conversion to Exudative AMD

Logistic regression models were trained on different feature sets in order to predict future conversion to exudative AMD. This analysis was limited to OCT volumes of eyes which did not already exhibit exudative AMD (2615 eyes, 1807 patients). For patients who developed exudative AMD, the earliest appearance of the corresponding ICD-10 code was recorded as the conversion date.

We applied logistic regression analyzes to predict future conversion to exudative wet AMD based on our extracted features. EHR and machine-read OCT B-scan features were used as input features to predict a future diagnosis of exudative Wet AMD. We compared four different combinations of feature groups: 1) the *current state* model used only the current AMD status and time to next visit (defined above); 2) the *risk factors* model used the EHR demographic and comorbid risk factors as well as the time to the next visit; 3) the *biomarkers* model used *only* the machine-read OCT B-scan biomarkers, and 4) the *combined* model incorporated all of the features available. This analysis was repeated for time horizons ranging from three to 24 months.

Following the threshold optimization procedures outlined in (Yim et al. 2020), two operating thresholds were determined such that the model was expected to achieve 80% sensitivity and 90% specificity, respectively. Additionally, to assess how the model performs when they are optimized together instead of independently, we included a threshold for a balance of sensitivity and specificity by finding a threshold which maximizes true positive rate while minimizes false positive rate, i.e. finding a point on the ROC curve close to the top left corner.

We acquired performance metrics in the following manner: in one round of cross-validation we split the data set to train- and validation sets with a ratio of 7:1 in a way that the two sets were disjoint on the patient-level. The logistic regression model was trained on the train set, after which it was used to generate predictions on the same train set. Based on the performance metrics of this prediction, 3 operating thresholds (balanced, high sensitivity, high specificity) were determined. Then the trained model generated predictions between 0 and 1 for the validation set, and predictions were binarized according to the thresholds. From the binarized predictions the rest of the performance metrics could be calculated.

Validations were performed for eight rounds (i.e. 8-fold cross validation) and the three operating thresholds were calculated as the means of the thresholds determined during the cross-validation.

95% confidence intervals were generated by running the described validation pipeline 125 times. Each time, 2000 samples were randomly selected from the full data set with replacement, on which the validation pipeline was executed yielding 125*8=1000 separate validation epochs.

#### Large scale validation of machine-read OCT features for diagnosis

As in the prediction task, the logistic regression framework was applied to diagnose the current exudative AMD status of each OCT B-scan volume (14615 OCT volumes, 4182 patients). In this analysis, two feature sets were compared: (1) EHR-derived risk factors (age, Smoking Status, Race, Ethnicity, Sex, and Chronic comorbidities), and (2) EHR-derived risk factors and machine-read structural OCT B-scan AMD risk factors (hcDVh, IHRF, hDC, SDD, and RPD). Model performance was quantified in terms of area under the receiver operating characteristic curve (AUROC) and area under the precision-recall curve (AUPRC).

## Data Availability

The data are not publicly available due to institutional data use policy and concerns about patient privacy. The data set consists of 14,615 OCT volumes and corresponding electronic health record data collected from 4,182 patients in 2018 at Doheny UCLA Eye Centers. In order to apply for data access, please visit https://doheny.org or reach out to.

https://github.com/Pairas92/AMD-prediction

https://doheny.org

## Other Acknowledgements

none

## Ethics Statement

The research was approved by the institutional review boards (IRBs) of the respective institutions and all subjects signed written informed consent. All research was conducted in accordance with the tenets set forth in the declaration of Helsinki. All imaging data were transferred to the Doheny Image Reading Center (DIRC) in a de-identified fashion. The image analysis research was approved by the UCLA IRB.

## Author Contributions

### Author Contributions

All Authors contributed to the conception and design, administrative support, provision of study material and patients, collection and assembly of data, data analysis and interpretation, manuscript writing and final approval of the manuscript. All authors read and approved the final manuscript.

## Competing Interests

### Financial Support/Fundings

None

### Financial Disclosures

Akos Rudas: none; Jeffrey N Chiang: none; Giulia Corradetti: none; Nadav Rakocz: none; Eran Halperin: United Health Group (C); SriniVas Sadda: Amgen (C), Allergan (C), Genentech-Roche (C), Oxurion (C), Novartis (C), Regeneron (C), Iveric (C), 4DMT (C), Centervue (C, S), Heidelberg (C, F, S), Optos (C, F, S), Carl Zeiss Meditec (F, S), Nidek (S), Topcon (S)

## Code Availability

The code used for the analysis is available on the following GitHub repository: https://github.com/Pairas92/AMD-prediction

## Notes

### Funding Statement

The author(s) received no specific funding for this work.

### Author Declarations

The Institutional Review Board of University of California Los Angeles (UCLA IRB; IRB #15000083AM00056 Ocular Imaging Study; Doheny UCLA Eye Centers, Pasadena, California) gave ethical approval for this work.

